# Using Machine learning to predict chronic kidney diseases among diabetic patients in Rwanda

**DOI:** 10.1101/2024.10.23.24315998

**Authors:** Rugamba Rugero Fiacre, Silas Majyambere, Baza Noella Confiance, Niyomugaba Germain, Uwera Aliane, Nemeyimana Patrick, Pierre Dukuziyaturemye

## Abstract

**Background:** Chronic Kidney Disease (CKD) is a significant complication in people with diabetes, leading to serious adverse health outcomes and increased healthcare costs globally individually and on healthcare systems. This problem become more complicated when it is in Low and middle in countries including Rwanda when access to early diagnostic services is limited. Early prediction and intervention can improve patient outcomes and reduce the burden on healthcare systems.

**Objective:** This study aimed to develop and evaluate a machine learning model for predicting CKD in diabetic patients, tailored to the Rwandan population, using Electronic Medical record Data.

**Methodology:** Secondary data were extracted from OpenClinic, an electronic medical record (EMR) system used at Kigali University Hospital, covering a period of 10 years from 2013 to 2023. The final cleaned dataset was used to train four machine-learning models: Logistic Regression (LR), Random Forest (RF), Decision Tree (DT), and Extra Gradient Boosting Machine (XGBoost). XGboost was noted as the best performer with the AUC score of 0.98 and accuracy of 95.67%.

**Results:** The findings revealed that XGBoost was highly effective in predicting chronic kidney disease, achieving an accuracy of 95.76% and an AUC score of 0.98. Given that the dataset was collected from the local population, this study confirms that machine learning algorithms can assist clinicians in Rwanda in diagnosing chronic kidney disease in its early stages.

**Conclusion:** This study demonstrates the potential of machine learning models in predicting chronic kidney disease (CKD) in diabetic patients, highlighting the importance of local datasets for optimizing model performance in specific populations. These findings suggest that machine learning can effectively assist existing medical techniques in the early diagnosis of CKD in Rwanda.

**Author summary:** In this study, we trained machine learning model to predict the risk of chronic kidney disease (CKD) in patients with diabetes, using a dataset collected in Rwanda. Early detection of CKD is crucial, as it allows healthcare providers to intervene sooner, improving patient outcomes, potentially reducing financial, and health burden on the patients. We processed the data, by handling different available data issues and statistically created new features such as anemia status and length of hospital stay to improve the model’s predictions. The final model, XGBoost provides insights that it can help health providers to identify high-risk patients and plan personalized care more effectively.

This study highlights how data-driven solutions can support healthcare delivery in resource-limited settings, by enhancing early diagnosis especially at primary healthcare level. By integrating this predictive tool into routine clinical workflows of Electronic Medical Record, healthcare institutions can make better clinical decisions that improve patient care and outcomes. This project contributes to the growing field of health informatics in Africa and shows the potential of applying advanced analytics to solve local health challenges.

## Introduction

While increasing attention is paid to the rising prevalence of chronic diseases in Africa, there is little focus on chronic kidney disease (CKD) though it directly resulted in an estimated 1.23 million deaths worldwide in 2017 and 1046 Deaths in Rwanda in the same year with an estimated prevalence of 10% of adults aged more that 18years old (1) (2). According to the International society of nephrology (ISN), worldwide in 2020 about 850Million People were suffering chronic kidney disease at different stages, and its prevalence has a rising trend (3). CKD is one of the leading causes of cardiovascular diseases, which are then the most common cause of death worldwide (4).

CKD is a progressive health condition where the kidneys gradually lose their ability to perform its functions properly. It affects people of all ages and all backgrounds but it is most common in older people and those with underlying medical conditions like diabetes and/or hypertension. For the people who has CKD, there is a decline in overall kidney function leading to losing the ability to filter out waste products and maintaining fluid and acid balance (5) .

As kidney progresses, it leads to more serious complications that needs critical medical attention and these complications affect multiple organ systems and significantly affect the overall status of health. CKD often causes or worsen blood pressure which further the damage on the kidneys and increase the risk of heart diseases like heart failure, coronary artery diseases and stroke (6).

Currently, in Rwanda to diagnose Chronic Kidney diseases, a urine examination is a must irrespective the level of the healthcare institution with the aim of dipstick test, microspic analysis and identifying the quantity of protein that the kidney is able to excrete (7). In microscopic analysis, the target to look for the presence of essential blood cells in the urine. The presence of Red Blood Cells (RBC) in the urine, known as hematuria, suggest glomerular damage, hence the cells leak in urine (8).

Instead of dipstick, Albumin – to-creatinine ratio (ACR), a more accurate technique is used in some cases as it is considered to be the best measure of proteinuria levels (9). It gives standardized measure of proteinuria making it easier to compare results across different settings with different equipment’s. It is also believed to be more sensitive than dipstick in detecting micro albuminuria which is an early sign of CKD (10). However, ACR can be influenced by the flow rate which result to not always accurately reflect proteinuria. Also, in cases with severe proteinuria, ACR is prone to failing to accurately reflect the total protein excretion (11).

eGFR is another test kidney functioning test. it is actually a measure to know how the kidney is able to filter waste from the blood in the process of making urine though it can be slightly affected by other conditions like dehydration or existing acid-base disorders (12). It is calculated based on blood creatinine levels, age, gender and in some cases race (13). High eGFR, >90ml, indicate well-functioning kidneys while low eGFR indicates severely damaged kidneys; when it is <15ml/kg the suffering patient have to depend only on renal replacement therapy like dialysis for lifetime or surgical kidney transplant (14).

While the mentioned tests are valuable for evaluating kidney function, they often fail to detect chronic kidney disease (CKD) in its early stages, when the kidneys are only minimally damaged. In these initial stages, the kidneys may still compensate for damage, leading to subtle changes in laboratory results that might not be apparent to measurement equipment (15). Additionally, factors such as age and gender can influence the interpretation of these results, increasing the risk of missed diagnoses (16).

With the rapid advancement of technology, clinical researchers, healthcare professionals, and ICT engineers are collaborating to explore the use of machine learning algorithms in healthcare settings, particularly for diagnosing non-communicable diseases including CKD, which often presents with no clinical symptoms at its early stage. Availability of electronic medical record (EMR) data is a great asset for not only healthcare administrators to help in administrative decision making, but also the health informaticians benefit this data for training ML algorithms that are able to assist in clinical decision making including early diagnoses (17).

Machine learning was proven as an important, evolving tool for accurately predicting the risk of CKD in patients with Diabetes, Hypertension, or other non-communicable diseases (18) (19). With relevant, well-prepared data, ML algorithms are able to learn relationships between risk factors of a disease so that it can forecast a trend for unseen data. For CKD, the main factors associated are age, hypertension, diabetes, Glycated Hemoglobin (Hb1Ac), serum creatinine levels, proteinuria, and Estimated Glomerular Filtration Late (eGFR), and others, from that, ML can predict CKD risk in new patients based on individual characteristics (20).

Machine learning has been widely used to predict CKD progression using algorithms like Random fore, support vector machines, and neural networks. A systematic review comparing these models demonstrated that RF and Gradient boosting to have excellent performance metrics results in handling non-linear data like eGFR and albuminuria levels (21) (22).

Also ML models have proven effectiveness in risk stratification of CKD patients which is crucial for interventions targeted on specific group. The study used XGBoost, Logistic Regression and decision trees to classify patients based on both clinical and demographic data. Here, XGBoost provided best performance with an accuracy of 92% and precision of 90% outperforming other models in identifying patients at risk of CKD (23).

A study applied SVM and RF to clinical datasets, using clinical features like Blood pressure, diabetes status and serum creatinine levels. RF achieved a promising highest accuracy of 93%. It was able to achieve this because of strength in managing high dimensional data, and proved its potential use in routine clinical practice (24).

Deep learning (DL) techniques like convolutional neural networks (CNN) and recurrent neural networks (RNN) are also great in disease diagnosis. These models were applied on a large a clinical dataset including time-series la results and showed better performance compared to previous studies which used standard ML algorithms. There was improvement of 5-10% improvement over standard algorithms (25).

## Methodology

In this research study, we utilized a comprehensive dataset extracted from the OpenClinic system of the University Teaching Hospital of Kigali. This dataset specifically focuses on patients diagnosed with diabetes who were managed at this facility. A range of clinical data, demographic information, and laboratory results were collected. The study retrospectively covers a period of ten years, from 2013 to 2023.

To train the model, the dataset was split in a 75:25 ratio, meaning that 75% of the entire dataset was used for model training, while the remaining 25% was reserved for model testing. Four machine learning classifiers—Random Forest, Extra Gradient Boosting (XGBoost), Decision Tree, and Logistic Regression—were trained and evaluated based on the Accuracy and Area Under the Curve (AUC) of the Receiver Operating Characteristic (ROC).

### Dataset Description

At the end of data collection, we compiled a dataset containing 6,900 instances and 29 features. Among these, 10 were numerical variables and 19 were categorical variables. To enhance the comprehensiveness of the dataset, five additional features were statistically engineered: anemia status, derived from hemoglobin levels; days of stay, representing the number of days the patient spent in the hospital; and medical insurance status, indicating whether the patient had insurance coverage. Additionally, 10 features that were not relevant to the model’s training were excluded, resulting in 24 feature**s** being retained. Each individual feature is illustrated in **Figure 1**.

**Figure 1.**
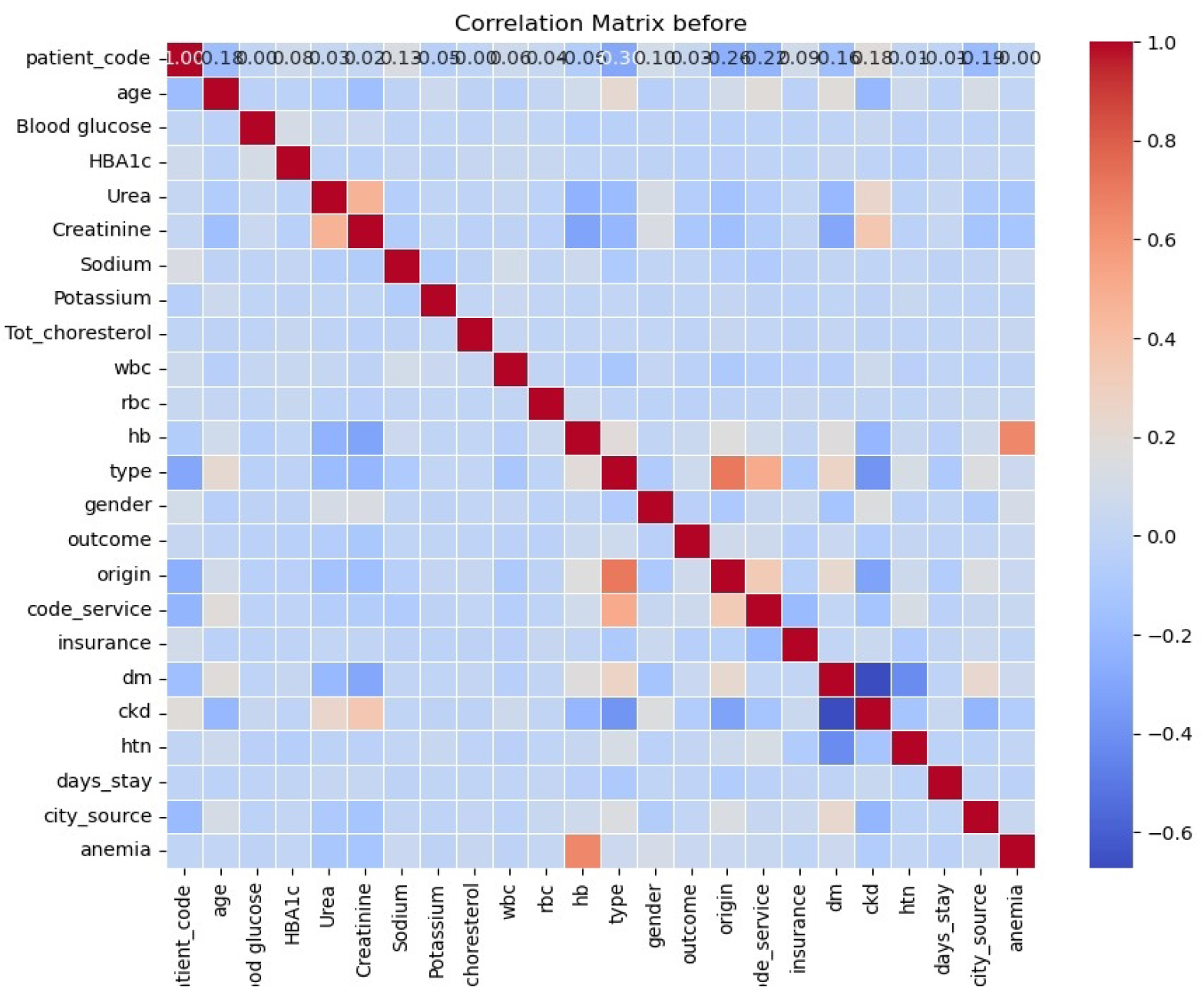
Correlational matrix of numerical features.

**Figure 2.**
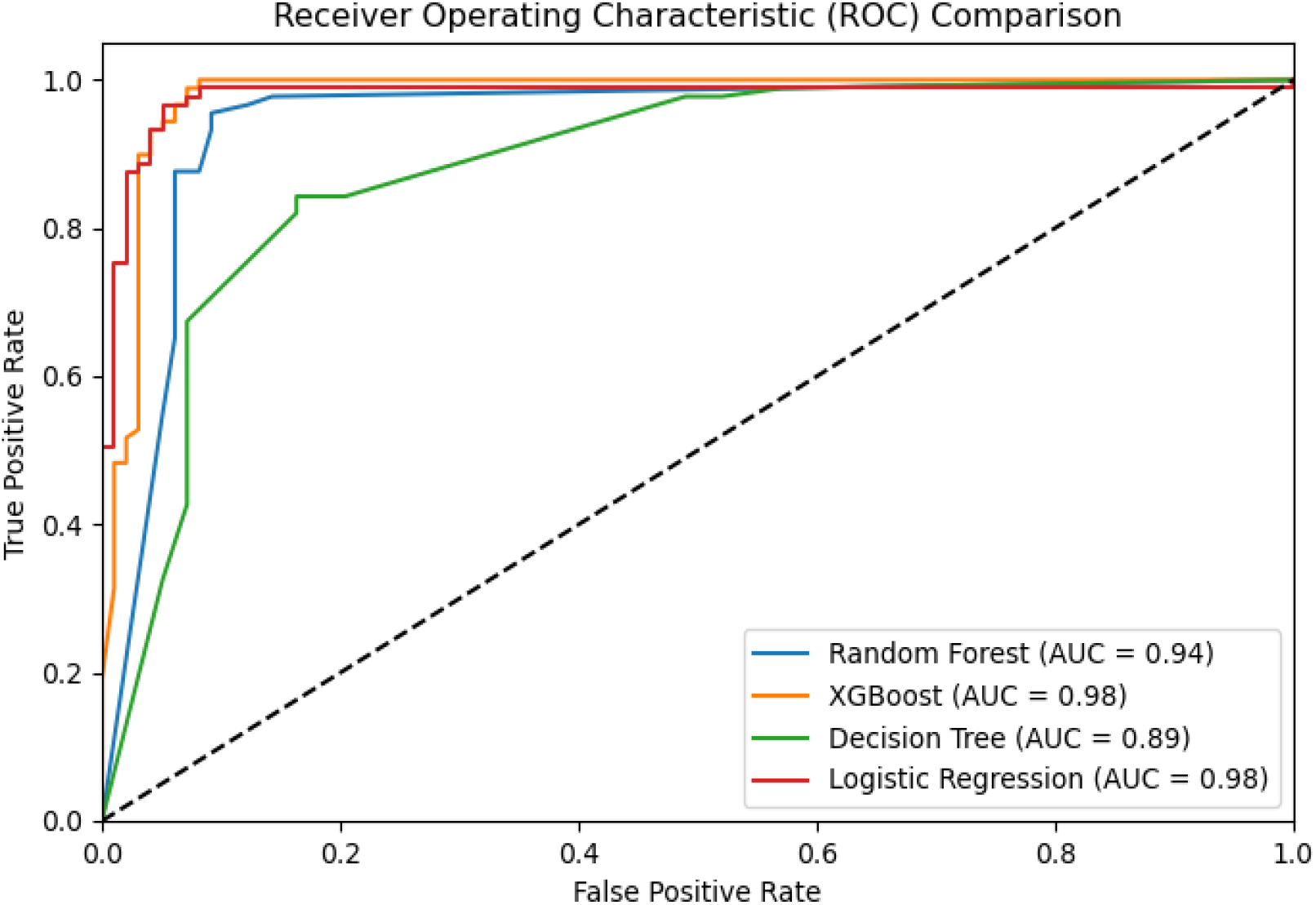
comparison of achieved AUC ROC results.

The dataset was thoroughly examined for data quality issues. No duplicates were identified, and no feature exhibited missing data exceeding 35%. To address the remaining missing values, the K-Nearest Neighbors (KNN) imputation method was employed, ensuring appropriate data filling. Additionally, proper data formatting was conducted to ensure correct interpretation by Python. An assessment for multicollinearity indicated there was minimal correlation issue, as illustrated in Figure 3. Grid Search was used to select and tune the best hyperparameters.

### Machine learning classifiers

#### Logistic regression

Logistic regression function is a function that was initially developed by Pierre Francois Verihulst, a Belgian Medical Doctor and Mathematician, in 1840s based on research on modelling population growth (26). Many subsequent researchers continued to work on it improving it until 1960 where it become how we see it today. Logistic regression is applied in different fields from social sciences, public health and medicine, to machine learning. This function is also used in engineering to predict the success or failure of a system or model (27).

Logistic regression models the probability of a binary outcome by mapping input variables to a value between 0 and 1 using the logistic (sigmoid) function. Its counterpart, linear regression predicts a continuous dependent variable based on a linear relationship between independent variables (27).

Mathematically Logistic regression for P: R → (0,1) can be represented as follows:

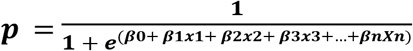

#### Random Forest

Random forest also known as a random decision forest is used mostly in classification and regression tasks that are built on combining different decision trees through out training time. For classification tasks like this study, the output of decision tree is the class that have been selected most times (28).

#### Decision Tree classifier

A decision tree is a visual and intuitive model used for both classification and regression tasks in machine learning. It represents decisions and their possible consequences in a tree-like structure, where each internal node signifies a feature or attribute, each branch represents a decision rule, and each leaf node represents an outcome or final decision. The model splits the dataset into subsets based on feature values, aiming to improve prediction accuracy with each split. Decision trees are favored for their simplicity, ease of interpretation, and ability to handle both numerical and categorical data (29).

**Gini impurity** is calculated for each split to evaluate the impurity of the resulting subsets:

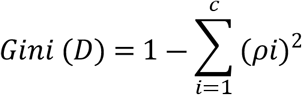

**Entropy** measures the uncertainty in the dataset and is another criterion used to evaluate splits:

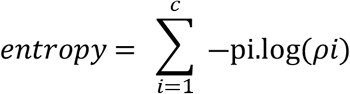

#### XGBoost

XGBoost (Extreme Gradient Boosting) is an advanced implementation of gradient boosting designed for speed and performance in machine learning tasks, particularly for classification and regression. It builds an ensemble of decision trees in a sequential manner, where each new tree corrects errors made by the previously built trees. The model minimizes a loss function, such as mean squared error for regression, and incorporates regularization to prevent overfitting. The core of XGBoost optimization process can be captured by its objective function:

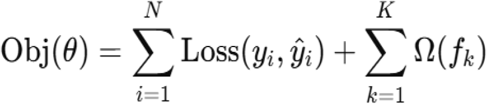

## Results and discussion

In this study, data issues were handled, hyperparameters tuned and 4 machine learning classifiers trained. Each algorithm used was described in methodology part, above.

To evaluate the performance of each model, a confusion matrix was generated. A confusion matrix is a structure like a table which is outputted to show the key performance metrics like: accuracy, Recall, Precision, F1-score and specificity.

In this study, True Positives (TP) refer to instances where the model correctly predicts the presence CKD. True Negatives (TN) indicate when the model accurately predicts its absence. False Positives (FP) occur when the model incorrectly identifies CKD when it is not present, potentially causing unnecessary anxiety. False Negatives (FN) happen when the model fails to detect CKD when it is actually present, risking untreated conditions (26).

To obtain the classification report, various constituent variables are used to calculate the parameters essential for determining the accuracy score. Recall, or True Positive Rate, commonly referred to as sensitivity in epidemiology, is calculated using the formula provided in Table 1. The precision/positive predictive value (PPV) is calculated using the formula in Table1, while F1 score also known as tradition f-measure is the balance mean between the sensitivity and precision (26). The detailed description of classification report got by this study from all trained models is described in Table 2.

**Table 1.** Description of the dataset.

| Description | Values |
| --- | --- |
| Data source | Kigali University teaching Hospital |
| Period | 2013 to 2023 |
| No of instances | 2670 |
| Number of features | 26 |
| Have CKD | 446 |
| Do not have CKD | 2204 |

**Table 2.**
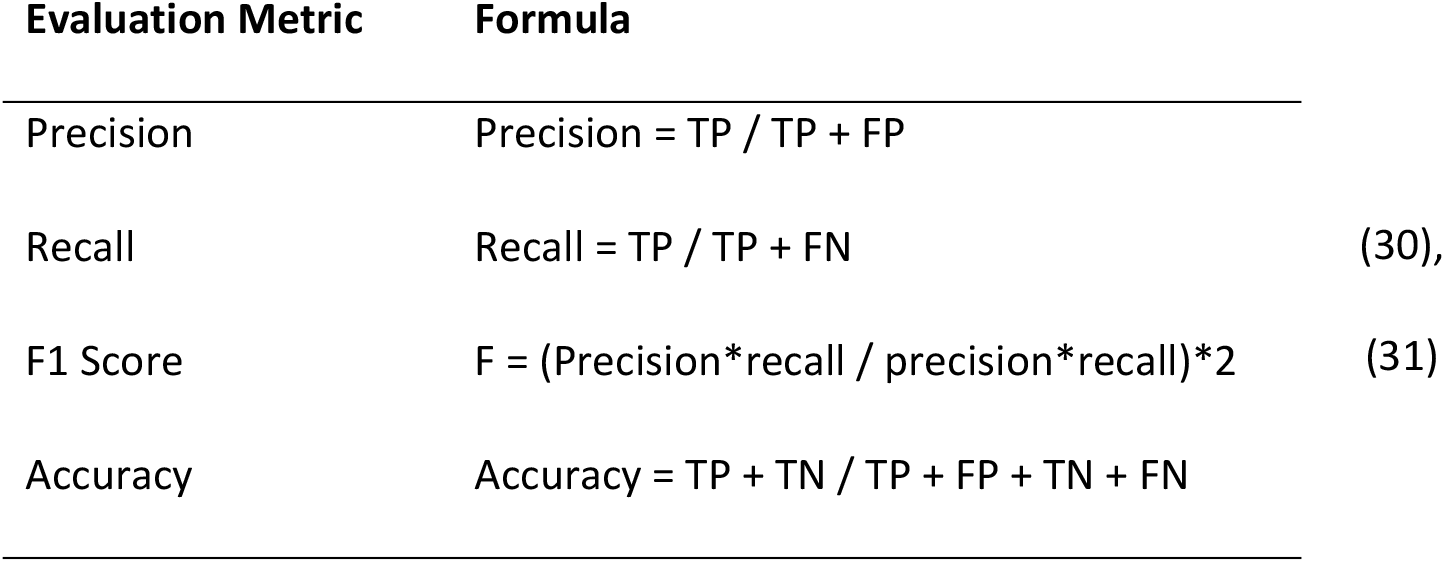
Confusion matrix metrics with formulas.

| Evaluation Metric | Formula |
| --- | --- |
| Precision | Precision = TP / TP + FP |
| Recall | Recall = TP / TP + FN (30), |
| F1 Score | F = (Precision*recall / precision*recall)*2 (31) |
| Accuracy | Accuracy = TP + TN / TP + FP + TN + FN |

**Table 3.**
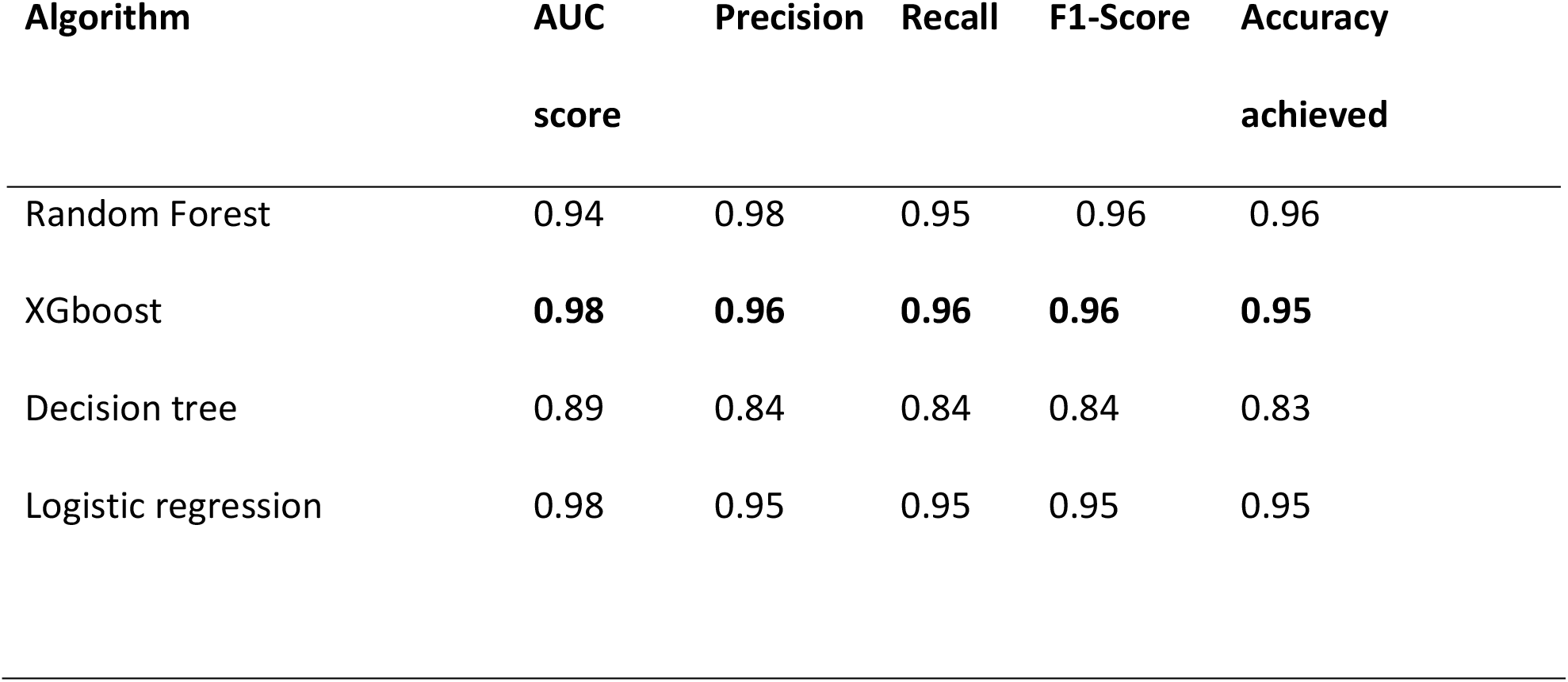
Classification results achieved by this study.

| Algorithm | AUC<br>score | Precision | Recall | F1-Score | Accuracy<br>achieved |
| --- | --- | --- | --- | --- | --- |
| Random Forest | 0.94 | 0.98 | 0.95 | 0.96 | 0.96 |
| XGboost | **0.98** | **0.96** | **0.96** | **0.96** | **0.95** |
| Decision tree | 0.89 | 0.84 | 0.84 | 0.84 | 0.83 |
| Logistic regression | 0.98 | 0.95 | 0.95 | 0.95 | 0.95 |

To further evaluate our models’ performance, we plotted the ROC curves and calculated the AUC scores, as shown in Figure 1. The results indicate that XGBoost and Logistic Regression achieved the highest AUC score of 0.98, while the Decision Tree model had the lowest AUC score of 0.89.

Upon comparing model performance, both Logistic Regression (LR) and XGBoost achieved the same AUC and accuracy scores. However, they differed in their F1 scores, with LR scoring 0.95 and XGBoost scoring 0.96. Given these results, XGBoost was selected as the best model due to its superior F1 score, indicating better balance between precision and recall.

**Table 4.**
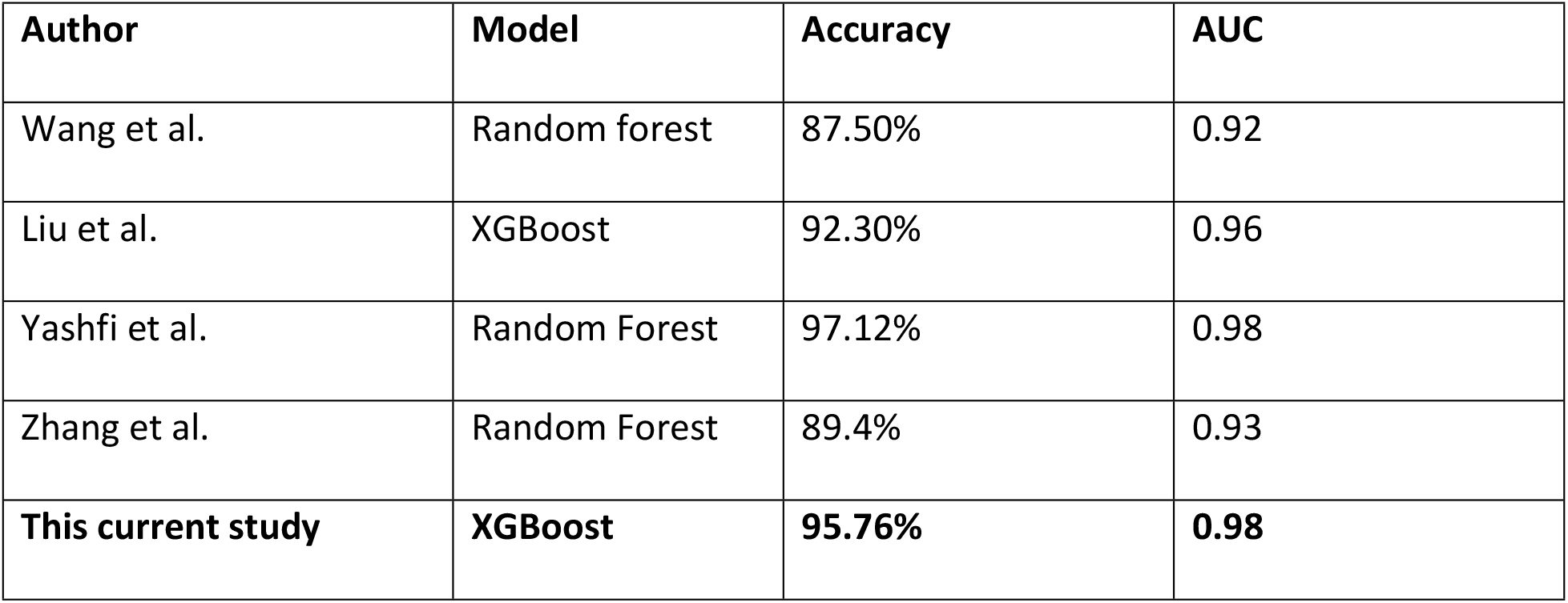
comparison of our results with previous studies.

| Author | Model | Accuracy | AUC |
| --- | --- | --- | --- |
| Wang et al. | Random forest | 87.50% | 0.92 |
| Liu et al. | XGBoost | 92.30% | 0.96 |
| Yashfi et al. | Random Forest | 97.12% | 0.98 |
| Zhang et al. | Random Forest | 89.4% | 0.93 |
| **This current study** | **XGBoost** | **95.76%** | **0.98** |

The results of our study demonstrate the efficacy of the XGBoost model in predicting chronic kidney disease, achieving an accuracy of 95.76% and an AUC of 0.98. These findings position our model competitively within the existing literature.

When comparing our results to those of previous studies, we note that Yashfi et al. reported a remarkable accuracy of 97.12% with a Random Forest model and an AUC of 0.98 (27) . While our accuracy is slightly lower, the high AUC indicates that both models are similarly effective in distinguishing between classes. Liu et al. also achieved a commendable accuracy of 92.30% with XGBoost and an AUC of 0.96, reinforcing the strength of this algorithm in this context (28).

In contrast, Wang et al. and Zhang et al. reported lower accuracy rates of 87.50% and 89.4%, with corresponding AUC values of 0.92 and 0.93, respectively (29) (30). These findings suggest that while Random Forest can be effective, our XGBoost model outperforms these earlier results, demonstrating enhanced predictive capability.

The consistent high performance of XGBoost across studies highlights its robustness as a machine learning tool for medical predictions, particularly for chronic kidney disease.

Additionally, the lack of statistically significant differences in accuracy between our results and those of Yashfi et al. underscores the reliability of our findings and their potential applicability in clinical settings.

The promising results of this study proves the transformative potential of machine learning in the diagnosis of chronic kidney diseases. As healthcare currently adopting advanced analytics, this developed model, XGBoost can significantly enhance early diagnostic accuracy and efficiency when it is deployed in EMR systems. This capability not only helps in early stages diseases detection but also supports personalized treatment strategies prescribed to individual patient profiles. The scalability of XGBoost further facilitates its implementation in diverse clinical settings, improving patient outcomes and optimizing resource allocation in healthcare systems. Using ML in CKD diagnosis presents an opportunity to revolutionize patient care, making it more proactive and data-driven.

### Limitations

This study was limited by its failure to identify and analyze chemical biomarkers in the blood of CKD patients. Research has consistently demonstrated the significance of these biomarkers in understanding disease progression and developing targeted treatments. Additionally, the study did not investigate genetic predisposition, which also play role in developing CKD.

## Conclusion

In this study, we utilized a dataset from the University Teaching Hospital of Kigali’s electronic medical records (EMR), which included a range of demographic and clinical variables. The dataset comprised various data types, including categorical variables. From this data, we developed a predictive model for chronic kidney disease using four machine learning algorithms: Random Forest, XGBoost, Decision Tree, and Logistic Regression.

To evaluate the models’ performance on unseen data, we generated a classification report and calculated the AUC. These metrics allowed us to assess the accuracy, precision, recall, and overall effectiveness of each model in predicting chronic kidney disease. XGBoost was noted as the best performing model with AUC, accuracy, F1score of 0.98, 0.95, 0.96 respectively. In contrast, the Decision Tree model exhibited lower performance, with an AUC of 0.89, an accuracy of 0.83, and an F1 score of 0.84. These results highlight the superiority of XGBoost in effectively predicting chronic kidney disease compared to the other algorithms tested. By applying this in clinical settings, it can positively impact patients’ health and assist medical practitioners to diagnose chronic kidney diseases in its early stages.

### Recommendations

To enhance the predictive power of the models developed in this study, it is recommended to increase the number of clinical features included in the analysis. By incorporating additional relevant variables—such as biochemical markers of CKD, and lifestyle factors—the model’s ability to capture the complexity of chronic kidney disease.

Additionally, exploring ensemble techniques could further boost model performance. Combining predictions from multiple algorithms, such as Random Forest and XGBoost, could leverage the strengths of each approach and mitigate individual model weaknesses. Future studies should focus on this point, which are crucial to improve predictive modelling in healthcare.

## Data Availability

The dataset used to train the model in this study is available in the main author Github's repository. No identification information is included in this dataset to ensure the privacy of the subjects involved

https://github.com/rug997/Masters-Thesis/blob/main/CKD%20Analysis.ipynb

## Acknowledgement

The authors would like to express their gratitude to the Centre of Excellence in Biomedical Engineering and e-Health at the University of Rwanda for providing access to educational resources and continuous support throughout this project

## Notes

### Competing Interest Statement

The authors have declared no competing interest.

### Clinical Trial

NA

### Funding Statement

The author(s) received no specific funding for this work.

### Author Declarations

The IRB of University of Rwanda, College of Medicine and Health Sciences issued ethical clearance. Additionally, the Ethical committee at University Teaching Hospital clearance to access data.

## References

1. Purcel, Carmel, Smith, Mali and Vos, Theos. Global, regional, and national burden of chronic kidney disease, 1990–2017: a systematic analysis for the Global Burden of Disease Study 2017. Washington : The Lancet, 2020.

2. FDA, Rwanda. Fourth Health sector strategic Plan. Kigali : Rwanda FDA, 2018.

3. Assessment of Global Kidney Health Care Status. Bello, Aki and Levin, A. 18, s.l. : JAMA Network, 2017, Vol. 385.

4. Chronic Kidney Disease and Cardiovascular Disease: Is there Any Relationship? Nathalie, Vallianou, Shah, Mitesh and Agathoniki, Gkogkou. 1, 2019, Vol. 15.

5. An Overview of Chronic Kidney Disease Pathophysiology: The Impact of Gut Dysbiosis and Oral Disease. Altamura, Serena, et al. 11, s.l. : Pubmed Central, 2023, Vol. 11.

6. Anupam, Agarwal and Karl, Nath. Pathophysiology of Chronic Kidney Disease Progression: Organ and Cellular Considerations. Chronic Renal Disease (Second Edition). s.l. : Science Direct, 2020.

7. Center, Rwanda Biomedical. National Strategy and Costed Action Plan for the Prevention and Control of Non-Communicable Diseases in Rwanda. Kigali : Rwanda Biomedical Center;, 2020.

8. Hematuria as a Risk factor for progression of chronic kidney disease and death: findings from the Chronic Renal Insufficiency Cohort (CRIC) Study. Orlandi, Paula F., et al. 9, s.l. : BMC Nephrology, 2018, Vol. 32.

9. Urinary albumin-to-creatinine ratio levels are associated with subclinical atherosclerosis and predict CVD events and all-cause deaths: a prospective analysis. Liu, Shanshan, et al. 3, s.l. : PubMed Central, 2021, Vol. 11. e040890.

10. Diagnostic accuracy of urine dipstick testing for albumin-to-creatinine ratio and albuminuria: A systematic review and meta-analysis. Mejia, Jhonatan R., et al. 11, Heliyon : s.n., 2021, Vol. 7. PMC8571083.

11. Albuminuria, Proteinuria, and Urinary Albumin to Protein Ratio in Chronic Kidney Disease. Wu, Men-Tai, et al. 2, s.l. : Journal of Clinical Laboratory analysis, 2012, Vol. 26. 10.1002/jcla.21487.

12. The Glomerular Filtration Rate: From the Diagnosis of Kidney Function to a Public Health Tool. Cusumano, Ana Maria, Tzanno-Martins, Carmen and Rosa-Diez, Guillermo Javier. s.l. : Frontier in Medicine, 2021, Vol. 8. 10.3389/fmed.2021.769335.

13. Measured and estimated glomerular filtration rate: current status and future directions. Levey, Andrew S, et al. 1, s.l. : PubMed, 2020, Vol. 16. 10.1038/s41581-019-0191-y..

14. Comprehensive review of current management guidelines of chronic kidney disease. Elendu, Chukwuka, et al. 23, s.l. : Wolters Kruwer Medicine, 2023, Vol. 102. 10.1097/MD.0000000000033984.

15. How Do Kidneys Adapt to a Deficit or Loss in Nephron Number? Fattah, Hadi, Layton, Anita and Vallon, Volker. 3, s.l. : American Physiology Society, 2019, Vol. 34. 10.1152/physiol.00052.2018.

16. Socioeconomic Status and Access to Healthcare: Interrelated Drivers for Healthy Aging. McMaughan, Darcy Jones, Oloruntoba, Oluyomi and Smith, Matthew Lee. 230, s.l. : Frontiers in Public Health, 2020, Vol. 8. 10.3389/fpubh.2020.00231.

17. Applications of Machine Learning Using Electronic Medical Records in clinical medicine. Schwartz, John T., et al. 4, s.l. : PubmedCentral, 2019, Vol. 16. PMC6945000.

18. Chronic kidney disease prediction based on machine learning algorithms. Islam, Ariful, Majumder, Ziaul Hasan and Hussein, Alomgeer. s.l. : Elsevier Journal of Pathology Informatics, 2023, Vol. 14. 10.1016/j.jpi.2023.100189.

19. A comprehensive review of machine learning techniques on diabetes detection. Sharma, Toshita and Shahcor, Manan. 30, 2021, Vol. 4. 10.1186/s42492-021-00097-7.

20. Risk factors for chronic kidney disease: an update. Kazancioglu, Rumeyza. 4, 2013, Vol. 3. 10.1038/kisup.2013.79.

21. Predicting chronic kidney disease progression with artificial intelligence. Isaza-Ruget, Mario A., et al. 20, s.l. : BMC Nephrology, 2024, Vol. 148.

22. Chronic kidney disease prediction based on machine learning algorithms. Islam, Ariful, Majumder, Ziaul Hasan and Hussein, Alomgeer. s.l. : Elsevier Journal of Pathology informatics, 2023. 10.1016/j.jpi.2023.100189.

23. Prediction Model and Risk Stratification Tool for Survival in Patients With CKD. S, Alexander, et al. 2, s.l. : Elsevier Kidney International report, 2018, Vol. 3. 10.1016/j.ekir.2017.11.010.

24. Chronic kidney disease prediction based on machine learning algorithms. Islam, Ariful, Majumder, Ziaul Hasan and Hussein, Alomgeer. 8, Ariful Islam; Ziaul Hasan Majumder; Alomgeer Hussein : Elsevier Journal of Pathology informatics, Vol. 14. 10.1016/j.jpi.2023.100189.

25. Gene Prediction: Methods, Challenges, and Future Perspectives. Chen, Qiannan. 6, Zhejiang : British Biomedical Bulletin, 2023, Vol. 11. 10.36648/2347-5447.11.2.4.

26. Schober, Patrick and Vetter, Thomas R. Logistic Regression in Medical Research. s.l. : PubMed Central, 2021. pp. 365–366. 10.1213/ANE.0000000000005247.

27. Sperandei, Sandro. Understing Logistic regression. New Dheli : Biochemia Medica, 2014. pp. 12–18. 10.11613/BM.2014.003.

28. Breiman, Leo. Random Forests. s.l. : SpringerLink, 2001. pp. 5–32.

29. Decision tree methods: applications for classification and prediction. Song, Yan-Yan and Lu, Ying. 2, Shangai : Shangai Archive of Psychiatry, 2015, Vol. 27. 10.11919/j.issn.1002-0829.215044.

30. A Review of Evaluation Metrics in Machine Learning Algorithms. Naidu, Grien, Zuva, Tranos and sibanda, Elias. s.l. : Springer Link, 2023. 978-3-031-35313-0.

31. Performance Metrics (Error Measures) in Machine Learning Regression, Forecasting and. Botchkarev, Alexei. 5, Ontario : Semantic, 2023, Vol. 12. https://www.bing.com/ck/a?!&&p=2023dd9629d57b82JmltdHM9MTcyOTU1NTIwMCZpZ3VpZD0yY2Fm MjdjYi04YTExLTY5MDktMTNiNS0zNGNjOGJlZDY4NDImaW5zaWQ9NTE5MQ&ptn=3&ver=2&hsh=3&fclid=2caf27cb-8a11-6909-13b5-34cc8bed6842&psq=Performance+Metrics+(Error+Measures)+in+Machine+.

32. Classification Techniques in Machine Learning: Applications and Issue. Soofi, Aized Amin. 76, s.l. : Journal of Basic & Applied Sciences, 2017, Vol. 13. 10.6000/1927-5129.2017.13.76.

33. Risk Prediction Of Chronic Kidney Disease Using Machine Learning Algorithms. Yashfi, Shanila Yunus, Islam, Ashikul and Pritilata. s.l. : Researchgate, 2020.

34. Long-Term Performance Prediction Framework based on XGBoost Decision Tree for Pultruded FRP Composites exposed to Water, Humidity and Alkaline Solution. Liu, Xing, Liu, TianQiao and Feng, Peng. s.l. : 284, 2022, Vol. 5.

35. Molecular contrastive learning of representations via graph neural networks. Wang, Yuyang, et al. s.l. : Nature Machin Learning, 2022, Vol. 4.

36. Artificial intelligence in recommender systems. Zhang, Qian, Lu, Jie and Jin, Yaochu. 12, s.l. : COmplex and Intelligent systems, 2020, Vol. 7.

